# Diabetes related antibody signature highlights ZnT8 as a novel antigenic target in rheumatoid arthritis

**DOI:** 10.64898/2026.09.25.26364002

**Authors:** Malin C. Erlandsson, Venkataragavan Chandrasekaran, Elena Neumann, Yuang Zhang, Malak Y.H. Alsoofi, Sofia T. Silfverswärd, Cristina Maglio, Ulf Muller-Ladner, Rille Pullerits, Maria I. Bokarewa

## Abstract

**Objective:** Autoantibodies are central for pathology of rheumatoid arthritis (RA) and diabetes mellitus (DM) carrying high predictive and diagnostic value. Here, we explore the frequency and functional potential of DM-related antibodies (DMab) in RA.

**Methods:** DMab targeting glutamic acid decarboxylase (GAD65ab), tyrosine phosphatase-like islet antigen 2 (IA2ab) and zinc transporter 8 (ZnT8ab) were measured in 326 DM-free RA patients (260 females, 66 males), using specific ELISAs. The DMab profile was analysed in relation to 30 clinical and serological variables. ZnT8^+^ cells were detailed by a public single-cell transcriptome in RA synovial tissue.

**Results:** DMab production was more frequent in females, dominated by ZnT8ab (86%), followed by GAD65ab (26.4%) and IA2ab (7.0%). DMab-positive patients, and specifically ZnT8ab-positive women, had higher levels of insulin and HOMA-IR index and lower serum IL-6, IFNγ, and IL-9. Multivariable logistic regression estimating ZnT8ab, IA2ab, GAD65ab production revealed that BDNF, insulin and DAS28 were positive predictors of ZnT8ab. Increasing ZnT8ab levels were inversely related to RAab in the mixed interaction model. CD14^+^ cell transcriptome identified association between an activated secretory cell type and increasing ZnT8ab levels. Additionally, ZnT8ab levels were in direct proportion with ZnT8 production in RA CD4^+^ T cells. Single-cell transcriptome of RA synovial tissue revealed ZnT8^+^ cells among the lubricin producing lining layer fibroblasts programmed for invasive behaviour.

**Conclusion:** This study reveals frequent production of ZnT8ab in non-diabetic RA patients and identifies their antigenic target expressed in lining synovial fibroblasts of RA joints.

## Introduction

Autoantibodies against self-proteins are a hallmark of autoimmune diseases and have significant diagnostic and predictive value in both rheumatoid arthritis (RA) and diabetes mellitus (DM).

The diabetes-associated antibodies (DMab) serve as key diagnostic markers and strong predictors for development of type 1 diabetes (T1D) and latent autoimmune diabetes in adults (LADA). The principal autoantigens include glutamic acid decarboxylase (GAD65), tyrosine phosphatase-like islet antigen 2 (IA2) and zinc transporter 8 (ZnT8), all of which are functionally linked with pancreatic beta-cells contributing to granule maturation, trafficking and insulin exocytosis.

The prevalence of individual DMab ranges from 24% to 65% in new-onset T1D ^1^ and from 16% to 34% in LADA ^2, 3^. In both cases, the DMab pattern is dominated by GAD65ab followed by IA2ab and ZnT8ab. Outside the pancreas, these proteins are involved in the regulation of neuroendocrine signalling molecules and synaptic activity and can contribute to neurological and other non-diabetic diseases.

DMab can also be detected in the healthy population ^4^. Increasing clinical evidence suggests that they may have diagnostic and prognostic value in non-diabetic conditions. High titres of GAD65ab are associated with neurological disorders including cerebellar ataxia, stiff-person syndrome and autoimmune epilepsy, often in the absence of DM. The antibodies are thought to inhibit GAD enzymatic activity, leading to reduced levels of gamma-aminobutyric acid in the brain and cerebrospinal fluid. The GAD65 antibody-mediated GABAergic dysfunction is now well recognized in the pathogenesis of stiff-person syndrome ^5^. Additionally, DMab, particularly GAD65 antibodies, are detected in 20-35% of patients with autoimmune thyroid disease, pernicious anaemia, celiac disease and autoimmune polyglandular syndromes, although their pathogenic role in these conditions remains unclear. ZnT8ab are found in about 24% of the non-diabetic first-degree relatives of T1D patients, in up to 16% of patients with autoimmune Addison’s disease ^6, 7^ and in around 31% of patients with autoimmune uveitis ^8^. Although a prognostic value of ZnT8ab has been acknowledged, no mechanistic role for ZnT8ab has been established in these diseases.

Similar to other autoimmune conditions, DMab can occur in RA but are not associated with DM development ^9^. During chronic inflammation, RA is associated with poor response to insulin signaling in the peripheral tissues, frequently resulting in hyperinsulinemia. Our recent studies demonstrated that long-lasting hyperinsulinemia changes immune responses through enhancing anergy and immunosenescence in CD4^+^ cells of RA patients ^10, 11^. It also increases histone acetylation thereby aiding survival and development of memory T cells ^10, 11^ and pointing at the increasing relevance of altered insulin signalling for RA pathology.

RA, a canonical autoimmune joint disease, is characterized by autoantibodies against the Fc-gamma region of IgG (rheumatoid factor, RF) and citrullinated proteins (ACPA), both detectable already at the preclinical stage of the disease ^12, 13^. These RA-specific antibodies are considered drivers of chronic joint inflammation and structural damage, marking severe and treatment-resistant RA. ACPA targets diverse citrullinated proteins, including vimentin, fibrinogen, enolase, fibronectin, filaggrin ^14, 15^. Antibodies against peptidylarginine deiminase (PAD) 4 and PAD2, the enzymes responsible for citrullination, the process converting arginine residues into the neutral amino acid citrulline, are associated with reduced disease severity suggesting their protective effect ^16, 17^. Notably, a subset of ACPA targeting citrullinated collagen exhibits experimentally confirmed anti-arthritic activity ^18^. Anti-collagen type II antibodies (CIIab) are frequently found in early RA ^19^. Although, CIIab, RF and ACPA are seldom measured together, they may share a convergent pathogenic mechanism involving cytokine production and early skeletal damage ^20^.

In recent years, antibodies against carbamylated (homocitrullinated) proteins have attracted attention, particularly in RF/ACPA-negative RA, suggesting alternative disease mechanisms ^21,22^. Experimentally, protein carbamylation may prime joint-directed immune responses ^23^. RA33 antibodies targeting the nuclear ribonucleoprotein A2/B1 are found in up to 35% of RA patients. They show no correlation to ACPA and erosive joint disease, aiding diagnosis in undifferentiated and seronegative RA ^24, 25^. Their association with milder RA suggests potentially protective properties. The palette of clinically relevant autoantibodies in RA is still growing.

Guided by an appealing hypothesis that the body produces both harmful and protective antibodies, we sought to identify specific DMab targets and whether they may contribute to RA molecular pathogenesis. Tissue expression patterns of autoantigens may provide new insights into their relationship with the autoimmune process and its pathological manifestations. In this study, we explored the frequency and clinical variables associated with DM-related antibodies against GAD65, IA2, and ZnT8 in non-diabetic RA patients. To assess functional importance of DMab, we analysed the specificity of CD4^+^ and CD14^+^ cell transcriptomes in relation to DMab levels. Finally, we explored expression of DMab target ZnT8 at a single-cell resolution in RA synovial tissue.

## Material and methods

### Study design and patient population

This longitudinal observational study included 326 patients with established RA enrolled between 2011 and 2019 at the Rheumatology Clinic, Sahlgrenska University Hospital, Gothenburg and the Northern Älvsborg County Hospital, Uddevalla. The clinical characteristics of the patients at inclusion are shown in **Table 1**.

**Table 1.** Clinical and demographic characteristics of the whole RA patient cohort and female RA patients.

| n=326 | DMab-negative<br>n= 147 | DMab-positive<br>n= 179 | p-values |
| --- | --- | --- | --- |
| Age, years | 55.2 ± 11.5 | 52.7 ± 12.6 | 0.12 |
| F/M, n | 93/55 | 167 /11 | p<10<sup>-8</sup> |
| BMI, kg/m2 | 26.5 ± 4.2 | 25.8 ± 4.7 | 0.032 |
| Smoking, current, n | 40 (27%) | 32 (18%) | 0.65 |
| DD, years | 12.0 ± 9.5 | 11.1 ± 8.8 | 0.36 |
| <3y, n | 28 | 32 | 0.79 |
| >10y, n | 66 | 69 | 0.24 |
| DAS28 | 2.74 ± 1.25 | 3.08 ± 1.37 | 0.059 |
| RAab, n | 128 (87%) | 158 (89%) | 0.74 |
| RF, n | 117 | 134 | 0.31 |
| ACPA, n | 104 | 114 | 0.18 |
| RF+ACPA, n | 93 | 90 | 0.11 |
| DMab | 0 |  |  |
| GAD65ab, positive |  | 55 (31%) |  |
| IA2ab, positive |  | 10 (5.6%) |  |
| ZnT8ab, positive |  | 157 (88%) |  |
| Two DMab |  | 43 (24%) |  |
| Oral corticosteroids, n | 15 (10%) | 25/179 (14%) | 0.31 |

DMab, diabetes antibodies; F/M, female/male; BMI, body mass index; DD, disease duration; DAS28, Disease activity score 28 joints; RAab, rheumatoid arthritis specific antibodies; RF, rheumatoid factor; ACPA, antibodies to cyclic citrullinated peptides; GAD65ab, antibodies to glutamic acid decarboxylase 65; IA2ab, antibodies to tyrosine phosphatase-like protein; ZnT8ab, antibodies to zink transporter 8. p-values are obtained by Mann-Whitney U test and chi-square test.

At inclusion, the standardized physical examinations by rheumatologist were performed in all RA patients and clinical and laboratory disease parameters were collected. To be included, the participants needed to fulfil the American Rheumatism Association 2010 revised criteria for RA ^26^. Exclusion criteria were serious physical or psychiatric illnesses that would hinder participation in either the physical examination, blood sampling or the patient interview, which was performed in Swedish. In this *at hoc* analysis, patients with diagnosis of diabetes Type 2 and Type 1 were excluded. At inclusion, all patients underwent a clinical assessment of disease activity in 28 joints and completed the Stanford Health Assessment Questionnaire disability index (HAQ) ^27^ Disease activity score was calculated based on examination of 28 tender and swollen joints (DAS28) and erythrocyte sedimentation rate (ESR). DAS28 below 2.6 indicated remission ^28^. Anti-rheumatic treatment at inclusion is shown in **Table 2**.

**Table 2.** Combinations of anti-rheumatic drugs in the studied RA patients.

| All, n=326 | Group size, n | MTX | TNFi | Other bDMARDs<sup>1</sup> | Other cDMARDs<sup>2</sup> | JAKi |
| --- | --- | --- | --- | --- | --- | --- |
| MTX | 265 | **135** | 63 | 35 | 21 | 11 |
| TNFi | 77 | 63 | **13** | 0 | 1 | 0 |
| other bDMARDs<sup>1</sup> | 54 | 35 | 0 | **12** | 2 | 6 |
| other cDMARDs<sup>2</sup> | 31 | 21 | 1 | 2 | **3** | 4 |
| JAKi | 25 | 11 | 0 | 5 | 4 | **5** |

<sup>1</sup>Other biological disease modifying anti-rheumatic drugs, bDMARDs: rituximab, n=14; tofacitinib, n=28; abatacept, n=12. <sup>2</sup>Other conventional (c)DMARDs: sulfasalazine, n=27; cyclosporine, n=3; azathioprine, n=1

### Ethics

The study was approved by the Swedish Ethical Review Authority (Dnr 659-11) and was conducted in accordance with national regulations and the International Conference on Harmonization Good Clinical Practice requirements, based on the Declaration of Helsinki. All patients provided written informed consent before being subjected to any study-related procedures. The study is registered at Clinical Trials.gov with ID NCT03449589.

Sample collection of the synovium and cartilage was approved by the local ethics committee of Region Västra Götaland (Dnr 2022-01977-02) and Justus Liebig University Giessen (66/08_74/05), and all patients gave written informed consent.

### Blood sampling and measurements

Blood samples were collected during the morning hours between 7 and 10 AM. The blood was obtained from the cubical vein into vacuum containers, and serum and plasma were stored at -70℃ until use.

### Isolation and stimulation of human CD4+ and CD14+ cells

Human peripheral blood mononuclear cells (PBMCs) from 56 non-diabetic RA patients were isolated from heparinized peripheral blood by density gradient separation on Lymphoprep (Axis-Shield PoC AS, Dundee, Scotland).

CD4^+^ T cells were isolated from PBMC by positive selection (11331D, Invitrogen, Waltham, MA, USA), and cultured at a cell density of 1.25 × 10^6^ cells/mL in wells coated with anti-CD3 antibody (0.5 microg/mL; OKT3, Sigma-Aldrich, St. Louis, MO, USA) in the Roswell Park Memorial Institute (RPMI1640) medium (Gibco, Waltham, MA, USA) supplemented with 5% foetal bovine serum (Sigma-Aldrich), 4 mM Glutamax (Gibco), 50 mM β2-mercaptoethanol (Gibco), and 50 mg/mL gentamycin (Sanofi-Aventis, Paris, France) at standard conditions of temperature, CO_2_ pressure, and humidity.

CD14⁺ cells were isolated from PBMC cultures by positive selection (kit #17858; StemCell Technologies, Vancouver, Canada) and propagated in the supplemented RPMI medium specified above at a concentration of 1.25 × 10⁶ cells/ml. Cells were stimulated with LPS (5 microg/ml) for 2 h. At the end of incubation, supernatants were collected for cytokine measurement, and cells were collected for transcriptional sequencing (RNAseq).

### Synovial fibroblasts

Synovial fibroblasts were isolated from either synovial tissue samples of RA patients after knee replacement surgeries at the Sahlgrenska University Hospital, Gothenburg (n=4) and the Agaplesion Markus Hospital, Frankfurt (n=4). Additionally, synovial fibroblast cultures were prepared from the knee synovial fluid (n=8) aspirated aseptically prior to intra-articular corticosteroid treatment. Synovial fluid was immediately transferred in heparinized vacuum tubes (Vacuette, Greiner Bio, Kremsmünster, Austria).

### Synovial fibroblast cultures

Experiments 1 and 2. Synovial tissue was digested (1 h Dispase-II-solution, 0.1 ml/ml, PAN-Biotech, Germany) ^29^ for fibroblast isolation. Cells were cultures up to passage 5 in DMEM (Paa Laboratories, Germany) containing 10% heat inactivated foetal calf serum (FCS, Sigma-Aldrich), 1 U/ml penicillin-streptomycin, and 1 mM HEPES (PAA Laboratories) at 37°C and 10% CO_2_.

Experiment 1. RASF were cultured for 48 h. The medium was replaced and cells stimulated with IL-1β (10 ng/ml; R&D) for 15 h ^30^.

Experiment 2. RASF were isolated from minced synovial tissue following treatment with 1 mg/ml collagenase/dispase (Roche, Mannheim, Germany) for 1 h at 37°C. Cells were cultured in growth medium containing DMEM medium (high glucose, GlutaMAX™ Supplement from Gibco™) supplemented with 10% heat-inactivated Gibco™ Foetal Bovine Serum, 50 mg/ml Gibco™ Gentamicin, and 100 U/ml Gibco™ Penicillin-Streptomycin at 37°C with 5% CO2. RASF (passage 6-7) from four patients were used in the experiment. In 6-well plates, 0.1 million cells were seeded and allowed to attach for 24 hours. Then the medium was replaced and cells were stimulated with TNF (10 ng/ml, R&D), insulin (25 nM, Humalog, Eli Lilly, Sweden AB) or TNF and insulin for 24h.

Experiment 3. From synovial fluid, cells were precipitated by centrifugation at 450g for 10 minutes, washed, and cultured in DMEM Glutamax (Gibco, Waltham, MA, USA) supplemented with 10% FCS (Sigma-Aldrich), 50 μg/ml gentamicin (Sanofi-Aventis, Paris, France) and 50 µg/ml normocin (Invivogen, San Diego, Ca, USA) at standard conditions of 37°C and 5% CO_2_. Treatment of RASF (passage 2-3) cultures was initiated after 48h. The medium was replaced and cells stimulated with TNF (10 ng/ml, R&D), insulin (25 nM) or TNF and insulin for 24h and 96h. Insulin was added daily.

At harvest, supernatants were collected and cells were lysed with the lysis buffer from the RNA extraction kit (see below) for further RNA preparation.

### Measurement of autoantibodies

DM autoantibodies were measured in serum according to the manufacturer’s instructions. The anti-glutamic acid decarboxylase IgG antibodies (GAD65ab) and anti-tyrosine phosphatase-like islet antigen 2 IgG antibodies (IA2ab) were measured using ELISA kits from Euroimmun® (Lübeck, Germany). The anti-zinc transporter 8 IgG antibodies (ZnT8ab) were analysed using an ELISA kit from RSR Limited (Cardiff, UK). The results are expressed in international units (IU/ml). Samples with the levels of DMab above 0.5 IU/mL for GAD65ab, 10 IU/mL for IA2ab, and 5 IU/mL for ZnT8ab were designated as positive. These cut-off levels are lower than the levels relevant for DM development ^4, 31^. The objective for using lower cut-off levels was to identify early and broader autoimmune activation that bears relevance for RA and systemic immune dysregulation.

Rheumatoid factor (RF) and anti-citrullinated protein antibodies (ACPA) were measured in serum samples at the accredited Laboratory of Clinical Immunology at Sahlgrenska University Hospital. ACPA was measured using an automated multiplex method (anti-CCP2, BioRad, Hercules, CA). Positive cut-off points were set according to the manufacturer at above 3.0 U/ml. Antibodies against Fc-region of gamma globulin (total RF) were measured by rate nephelometric technology (Beckman Immage 800, Beckman Coulter AB, Brea, CA) with a cut-off for RF positivity at 20 U/ml.

### Metabolic parameters

Sandwich ELISA kits were used to measure insulin levels (DY8056, R&D Systems, Minneapolis, MN, USA), plasma HbA1c (MBS2700040, MyBiosource, San Diego, CA, USA), and urine C-peptide levels were measured by ELISA (DICP00, RnD Systems), according to manufacturer’s instructions. Plasma glucose levels were measured using FreeStyle Lite kit (Abbott Diabetes Care Ltd., Oxon, UK) and used to calculate the homeostatic model assessment for insulin resistance (HOMA-IR) index. Total plasma cholesterol, HDL, LDL levels were measured at the accredited Laboratory of Clinical Chemistry at Sahlgrenska University Hospital. Body mass index (BMI) was calculated from body weight and height.

### Measurement of cytokines

The measurement of serum cytokines was performed using sandwich ELISAs (R&D Systems) for IL-4 (DY204), IL-8 (DY208), IL-9 (DY209) and IL10 (DY217B), VEGF (DY293B), BDNF (DY248) and FLT3-ligand (DY308). Serum IL-6 (M1916) and IFNg (M1933) were measured with a sandwich ELISA (Sanquin, Amsterdam, The Netherlands) as previously described ^32, 33^.

### Gene expression analysis by conventional RT-PCR

RNA from cultures was prepared using the Total RNA kit (Norgen, Ontario, Canada). Complementary DNA was prepared using High-Capacity cDNA Reverse Transcription Kit (Applied Biosystems, Foster city, CA). Quantitative PCR was performed with SYBR Green qPCR Mastermix (Qiagen) using a ViiA™ 7 Real-Time PCR System (Applied Biosystems). Melting curves for each PCR were performed between 60°C and 95°C to ensure specificity of the amplified product. The results are expressed in relative quantity (RQ) to the expression level in the control cells using the ddCt-method.

Primers were designed in-house using the Primer3 web client (https://primer3.ut.ee/) as described ^10^. Correct binding of primers was validated in UCSC In-Silico PCR (http://genome.ucsc.edu/cgi-bin/hgPcr) against the GRCh38/hg38 human genome assembly. Expression was measured using the following primers: *SLC30A8*, forward, CTTCTTGTCCCACCACTTGA, reverse, TGAAACAGTGCTGAGCGTCT; *IRS2*, forward, ATGATTGCAGAGGTCGTGGG, reverse, GGGAGGCTTCGATGACAACC. Expression was normalized to the reference gene *GAPDH* (TATAA Biocenter, Gothenburg, Sweden).

### Statistical analysis

Imputation of missing data (<5% for all parameters except DAS28 (16%) and ACPA (8%)) was performed using the non-parametric R package missForest (version 1.5). Raw data was log10-transformed to approximate normal distribution and z-standardised to achieve comparable beta coefficients.

To identify parameters associated with DMab production, an univariate logistic regression analysis adjusted by sex and age were performed in R studio using generalised linear model (glm). The variables with p<0.1 were included in the following multivariate logistic regression analysis with backward elimination performed in GraphPad Prism. The probability for DMab-positivity was estimated as area under the ROC curve (AUC) based on the variables that remained significant in the last step of elimination. For analysis of CD4^+^ and CD14^+^ cells transcriptome in relation to ZnT8ab level, a linear Lasso regression (Least Absolute Shrinkage and Selection Operator) model was used.

To assess the effect of combination of clinical parameters influencing the production of DMab, an interaction modelling analysis was performed controlling for age (Dmab_pos ∼ var1 * var2 + z_log_age), for sex and ZnT8ab (ZnT8_pos ∼ var1 + var2 + z_age + sex_female) and the likelihood-ratio test was used to test whether the interaction model performed better compared to the base model (Dmab_pos ∼ var1 + var2 + z_log_age).

Continuous raw data were analysed using non-parametric Mann-Whitney U test, or, in case of more than 2 groups, the Kruskal-Wallis test followed by Dunn’s post hoc test (Graphpad Prism). Correlations were tested using non-parametric Spearman (Graphpad Prism). All tests were two-tailed and P-values <0.05 were considered statistically significant.

### Transcriptome sequencing (RNA-seq)

Total RNA from CD4^+^ and CD14^+^ cells was prepared using the Norgen Total RNA kit (17200 Norgen Biotek, Ontario, Canada). Quality control was done by Bioanalyzer RNA6000 Pico on Agilent2100 (Agilent, Santa Clara, CA, USA). Deep sequencing was done by RNAseq (Hiseq2000, Illumina) at the core facility for Bioinformatics and Expression Analysis (Karolinska Institute, Huddinge, Sweden). Raw sequence data were obtained in Bcl-files and converted into fastq text format using the bcl2fastq program from Illumina.

### Bioinformatics

Transcriptome sequencing analysis was performed using the core Bioconductor packages in R-studio v. 4.4.1. Mapping of transcripts was done using Genome UCSC annotation set for hg38 human genome assembly. Using a design formula as ∼log_[ZnT8ab level]_scaled in the DESeq2 pipeline, we identified up regulated protein-coding genes with a base mean expression ≥1 that exhibited a significant change (|log₂FC| > 0.15, nominal *p* ≤ 0.05) per unit increase in serum ZnT8ab level. Analysis of Biological pathways and cellular compartment was done using the Gene Set Enrichment Analysis (GSEA) and Molecular Signature Database (MSigDB, Broad Institute, San Diego, USA.

SLC30A8-positive cells, defined as non-zero expression of *SLC30A8,* were identified in the single-cell transcriptome of synovial tissue of 82 RA patients (Synapse, syn52297840 ^34^), using the FetchData() function in Seurat.

A normalized expression of SLC30A8 was calculated in the lining synovial fibroblast cluster of RA synovial tissue based on model of the Human Protein Atlas. Normalized Counts Per Million (nCPM) in the context of the Human Protein Atlas were calculated by aggregating scRNA-seq reads across all cells within a specific cluster to create a pseudobulk profile within TMM-normalized (Trimmed Mean of M-values) protein-coding gene expression data.

## Results

### 1. The prevalence of DMab in non-diabetic RA patients

We found that 179 of 326 (54.9%) non-diabetic RA patients produced at least one type of DMab (**Figure 1A**). Clinical characteristics of DMab positive and negative RA patients are shown in **Table 1**. Among the DMab-positive patients, ZnT8ab were most prevalent, detected in 48.2% of the studied RA patients, followed by GAD65ab (16.9%) and IA2ab (3.07%). The level distribution of ZnT8ab, GAD65ab and IA2ab has a distinct pattern (**Figure 1B, 1C, 1D**). Using the canonical cut-off levels applied for assessment of risk for autoimmune DM for each DMabA, we found that 29 of 326 (8.9%) patients were positive, which is significantly higher than the expected 1-2% in healthy controls ^4, 31^ (**Figure 1E**). Additionally, a remarkable part of ZnT8 and GAD65 reactivity appeared in the borderline zone below the canonical cut-off level associated with DM but were still clearly above the top 75% of controls. Combination of two DMab was found in 43 of 179 patients (24%; 40f, 3m), where a combination of GAD65ab and ZnT8ab dominated (37/43, 86%).

**Figure 1.**
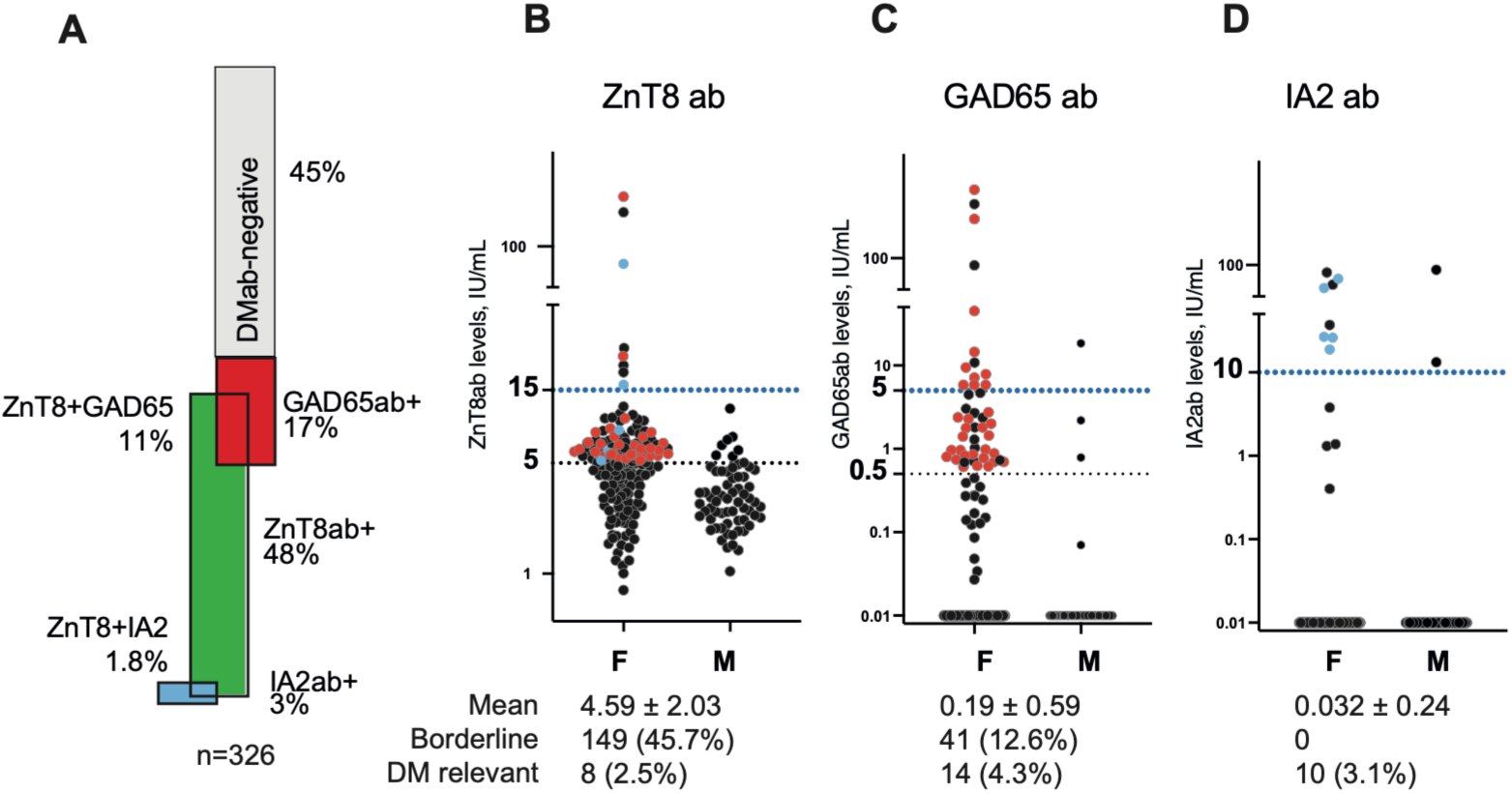
Diabetes mellitus related antibodies (DMab) in non-diabetic patients with rheumatoid arthritis (RA). A. Bar plot of DMab frequency in 326 patients. DMab were measured in serum by reactivity with zinc transporter 8 (ZnT8ab), glutamic acid decarboxylase (GAD65ab) and tyrosine phosphatase-like islet antigen 2 (IA2ab), using specific ELISAs. B, C, D. Dot plot of absolute antibody levels of ZnT8ab (B), GAD65ab (C), IA2ab (D) in female and male patients. Dotted horizontal line indicates the positive cut-off level and DM-relevant level for each antibody type. Red dots indicate patients positive for both GAD65ab and ZnT8ab, blue dots indicate patients positive for both ZnT8ab and IA2ab. DM relevant, ZnT8ab >15 IU/ml, GAD65ab >5 IU/ml, IA2ab >10 IU/ml. Any, 29/326, 8.9%. Mean excluding the DM-relevant patients. Boderline zone between the DM-relevant and cut-off used in this study, ZnT8ab, 5-15 IU/ml; GAD65ab, 0.5-5 UI/ml.

To explore, which of the variables were associated with DMab-positivity, we first applied univariate logistic regression analyses to 30 independent clinical and serological variables adjusting for sex and age (**Supplementary Table ST1A**). The variables with higher probability of relationship with DMab-positivity (p-values<0.2) were selected for the multivariate logistic regression analysis with backwards elimination. The variables that remained significant at the final step of the multivariate regression analysis included age, female sex, DAS28, and serum levels of insulin, IFNγ, and BDNF, as the main drivers of DMab production. The combination of these variables predicted DMab positivity with an accuracy of 0.783 (**Figure 2A**). The prediction strength by beta-estimate was comparable between the individual variables (**Figure 2B**). Next, we applied interaction modelling of the different variables postulating that a combination of these variables interferes with the prediction accuracy for DMab-positivity. Indeed, positivity for any DMab was significantly influenced by the combined effect of BDNF and smoking (beta=1.08, p=4.6e-4), and by serum levels of IL-8 (beta= -0.25, p=0.028, IL-9 (beta=0.22, p=0.085), and insulin (beta= -0.19, p=0.076) (**Figure 2E**). Together, these results demonstrated a frequent DMab production in non-diabetic RA patients, which was associated with both inflammation and anabolic variables.

**Figure 2.**
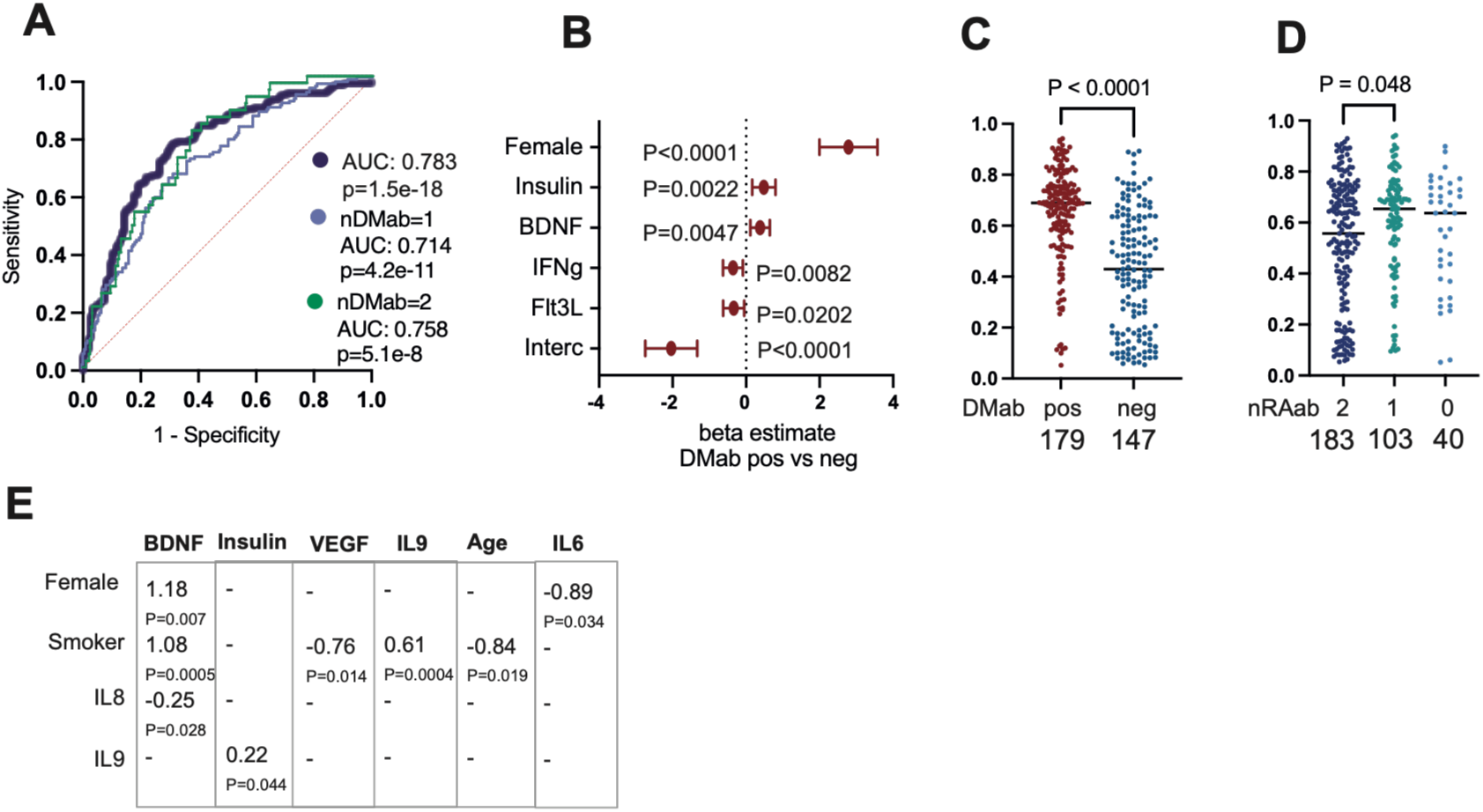
Serological associations of diabetes mellitus related antibodies (DMab) in patients with rheumatoid arthritis. DMab including ZnT8ab, GAD65ab, and IA2ab were measured in serum. In total, 175 of 326 patients were DMab positive. A. Receiver Operating Characteristic (ROC) curve of prediction for DMab positivity was calculated in the binary multivariate logistic regression analysis using presence of DMab as dependent to 30 serological and clinical variables. Area under the ROC curve (AUC) was calculated to estimate the accuracy of the model. B. Forest plot of beta estimates for the variables included in the DMab prediction model. Dots indicate beta estimate values, whiskers indicate 95% confidential interval. P-values are calculated by Wald statistics. C. Dot plot of individual prediction sum for each patient, based on the beta estimates shown in (B). Patients are grouped by presence of DMab (C) and by presence of rheumatoid arthritis specific antibodies (RAab) (D). P-values are calculated with non-parametric Mann-Whitney U test (C) or Kruskal-Wallis test with Dunns post hoc test (D). E. Table of beta interaction coefficients and likelihood ratio test P-values are shown in comparison to base model for DM ab positivity.

### 2. Production of DMab and RAab has different biological background

The combination of DMab and RAab was found in 158/326 (48%) patients (**Table 1**). Among those, 56.9% were positive for both RF and ACPA. Positivity for RAab was found equally frequent in DMab-positive as in DMab-negative group (89% vs 87%, respectively). Despite this seemingly equal distribution of RAab among the patients positive for ZnT8ab, GAD65ab and IA2ab (**Supplementary Table ST1**), the mixed interaction analysis revealed that ZnT8 levels were negatively associated with ACPA-positivity (estimate -1.84, adj.p=0.037) and the IA2ab levels with RF-positivity (estimate -3.14, adj.p=0.0013). Thus, presence of RF and ACPA conferred a reduced effect on DMab positivity, suggesting that a seronegative RA patient has a high likelihood of being positive for DMab. Consequently, we analysed if the estimated probability for DMab production separates the groups of RAab positive patients (**Figure 2C**). To investigate this, we summed up the estimated individual values for each patient and found that the probability of DMab production reduced with increasing production of RAab.

To identify molecular variables associated with RAab production, we performed the multivariate regression analysis using RAab-positivity as a dependent variable. This analysis revealed that increasing serum levels of IL-6 and survivin were positively associated with RAab, while current smoking, serum levels of IFNγ and VEGF were negatively associated with RAab production (AUC 0.748, p<0.001. **Supplementary Figure S1A, S1B**). The observed diversity of the predictor estimates related to DMab and RAab production strongly advocates for different biological mechanisms behind their production.

### 3. ZnT8 is the preferred antigen among DM-related antigenic targets in RA patients

Demographically, the DMab-positivity was identified frequently in female compared to male patients (64% vs 17%, p<10^-8^) (**Table 1**), with a strong over-representation in ZnT8ab. However, the high female preponderance was consistent for DMab of all three specificities. To adjust for the skewed gender distribution in further analysis, we investigated the female and male patients independently. Unexpectedly, we found no significant difference in the metabolic and inflammatory parameters between the male DMab-positive and DMab-negative groups (**Supplementary Figure S2A, S2B**) with the exception for the total cholesterol levels (p=0.039) and BMI (p=0.058) that tended to be lower in the DMab-positive males. Thus, we focused our further analysis on the females.

To investigate the relationship between serological and clinical variables and production of antigen-specific DMab, we performed a multivariable logistic regression analysis in females placing ZnT8ab, IA2ab, GAD65ab as binary dependent variables (**Figure 3 A, B, C**). We found that each subtype of DMab was significantly influenced by a particular set of variables individually associated with their production. Increasing levels of anabolic factors insulin and BDNF amplified the probability of ZnT8ab (**Figure 3A1, 3A2**); VEGF increased the probability of IA2ab (**Figure 3C1, 3C2**), while FLT3-ligand had an inverse effect on GAD65ab production (**Figure 3B1, 3B2**). Signs of active RA disease including the disease activity score DAS28 and the tender point count increased the probability of ZnT8ab production (**Figure 3A1, 3A2**), and ESR was associated with increased IA2ab production (**Figure 3C1, 3C2**). In contrary, pro-inflammatory cytokines reduced the probability of DMab; IFNγ had a negative impact on ZnT8ab production, IL-6 on GAD65ab, and IL-8 on IA2ab. Interaction modelling of the variables from the multivariate regression analyses confirmed the combinatorial impact of the identified parameters on specific DMab (**Figure 3A3, 3B3, 3C3**). Presence of GAD65ab together with active RA disease or serum insulin significantly influenced ZnT8 production, mirrored by the effect of BDNF together with IL-8, age, and smoking (**Figure 3A3**). Production of GAD65ab was dependent on the combined effect of FLT3L with IFNγ and HbA1c, and IL-6 with smoking respectively (**Figure 3B3**), while IA2 production was dependent on antibody presence of ACPA with VEGF, and ZnT8 presence with ESR and insulin (**Figure 3C3**).

**Figure 3.**
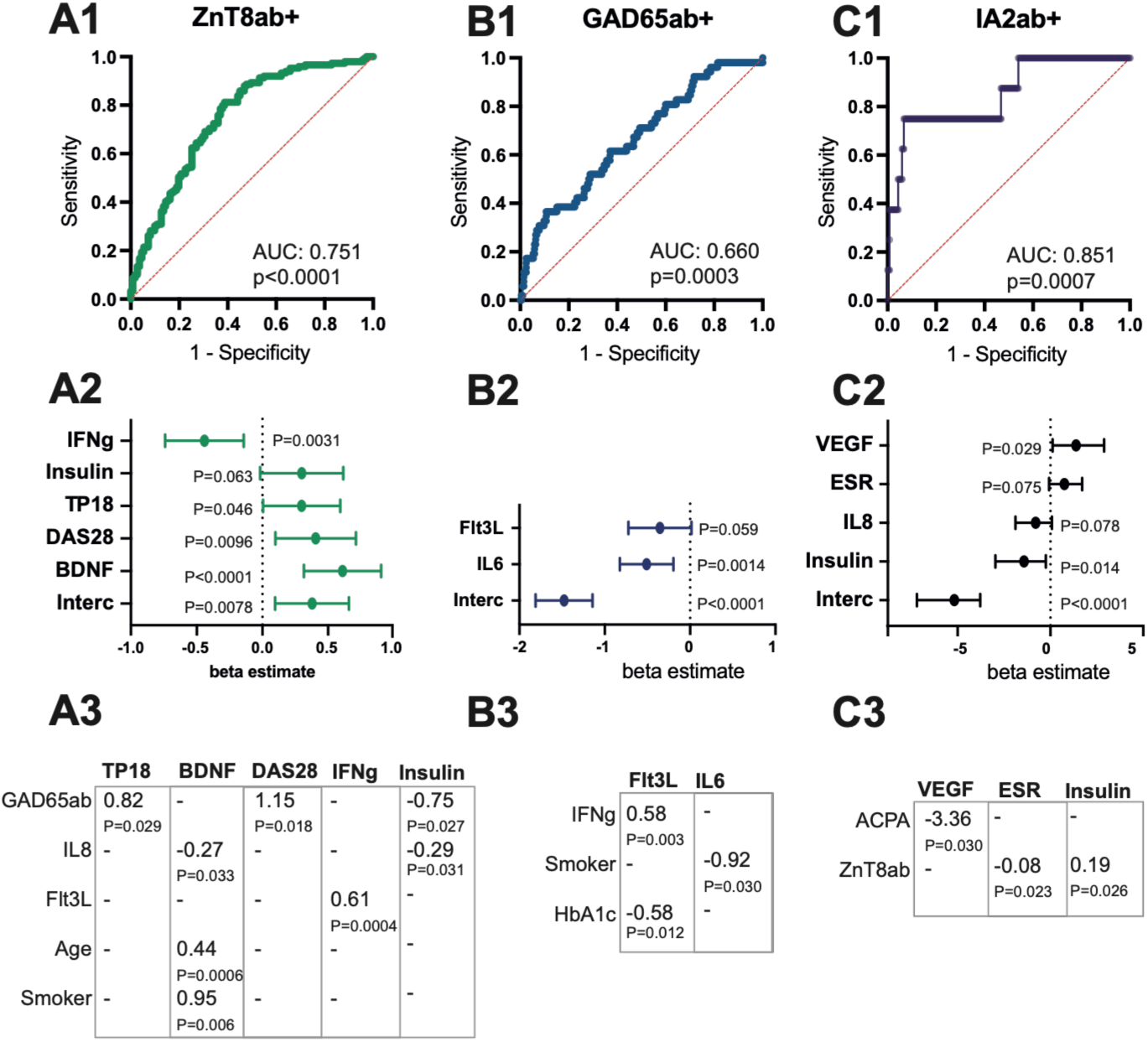
Serological association of different types of DM related antibodies in female non-diabetic patients with rheumatoid arthritis. Top row. Receiver Operating Characteristic (ROC) curve of prediction for ZnT8ab positivity (A1), GAD65ab positivity (B1) and IA2ab positivity (C1) was calculated in the binary multivariate logistic regression analysis using presence of antibodies as dependent to 30 serological and clinical variables. Area under the ROC curve (AUC) was calculated to estimate the accuracy of the model. Middle row. Forest plot of beta estimates for the variables included in the antibody prediction model for ZnT8ab (A2), GAD65ab (B2), and IA2ab (C2). Dots indicate beta estimate value, whiskers indicate 95% confidential interval. P-values are calculated by Wald statistics. Bottom row. Tables of beta interaction coefficients and likelihood ratio test P-values are shown in comparison to base model for ZnT8ab (A3), GAD65ab (B3), and IA2ab (C3) * P<0.05, ** P<0.01.

Among the female patients, DMab-positive group consisted almost exclusively of ZnT8ab-positive patients (95%, 149/157). The ZnT8ab-positive females had high levels of insulin (p=0.030) and IL-8 (p=0.043), and low levels of IL-6, IL-9, and IFNγ (**Figure 4**). No difference in HbA1c, C-peptide, BMI, age or total cholesterol levels was seen in females of any DMab specificity (**Supplementary Figure S3**). Among 40 females with a combination of ZnT8ab with either GAD65ab (n=37) or IA2ab (n=3), the presence of two DMab was associated with an additive effect on low serum levels of IL6 and IFNγ (**Supplementary Figure S3**), but not on the levels of IL-8, IL-9, nor insulin.

**Figure 4.**
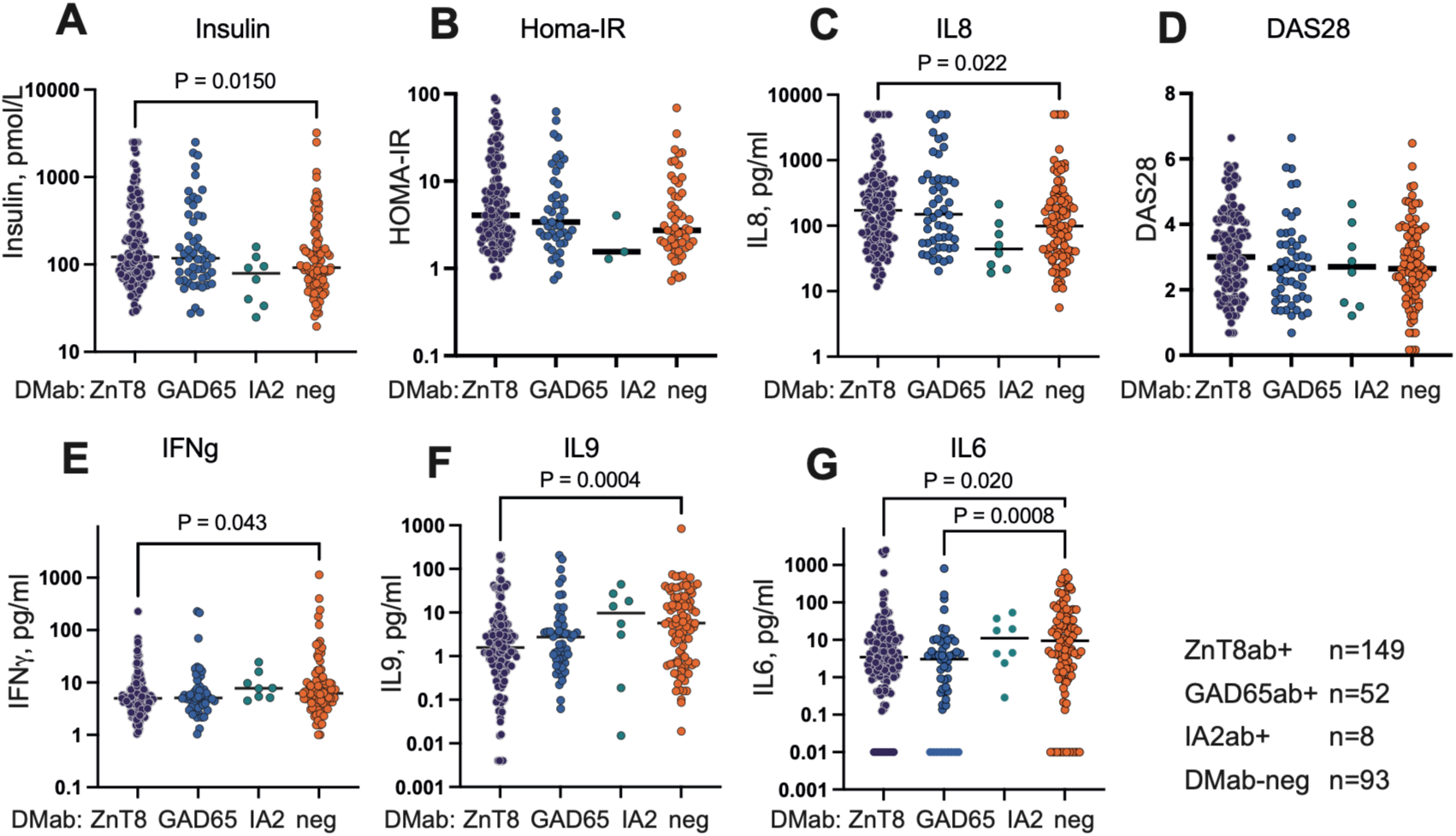
Serological characteristics of non-diabetic female patients with rheumatoid arthritis grouped by presence of diabetes mellitus related antibodies. Dot plot of clinical and serologic variables grouped by antibody type, ZnT8ab (n=149), GAD65ab (n=52), IA2ab (n=8), DMab-negative group (n=93). Patients with combination of autoantibodies are included in both groups. P-values are calculated against DMab negative group with non-parametric Mann-Whitney U test.

### 4. Connection between ZnT8ab levels and transcriptome of CD4^+^ and CD14^+^ cells

To investigate the cellular and molecular mechanisms associated with ZnT8ab production in more detail, we explored the transcriptome of CD4^+^ and CD14^+^ cells from 56 female RA patients in regression to their serum ZnT8ab levels. Unexpectedly, the transcriptome analysis of CD4^+^ cells revealed a positive relationship between the serum Znt8ab levels and the expression of *SLC30A8* mRNA coding for ZnT8 (log2FC 1.67, p=0.0037). Moreover, measurable expression of the *SLC30A8* gene in CD4^+^ cells was detectable with a normalized expression above 1, in 14% (8/56) of the patients (expression range 1.33-13.30). The patients with measurable *SLC30A8* expression in CD4^+^ T cells had higher levels of ZnT8ab in serum (**Figure 5A**). The pathway enrichment analysis of genes upregulated in CD4^+^ cells in regression to increasing ZnT8ab levels were different from those identified in CD14^+^ cells and reflected broad cell physiology.

**Figure 5.**
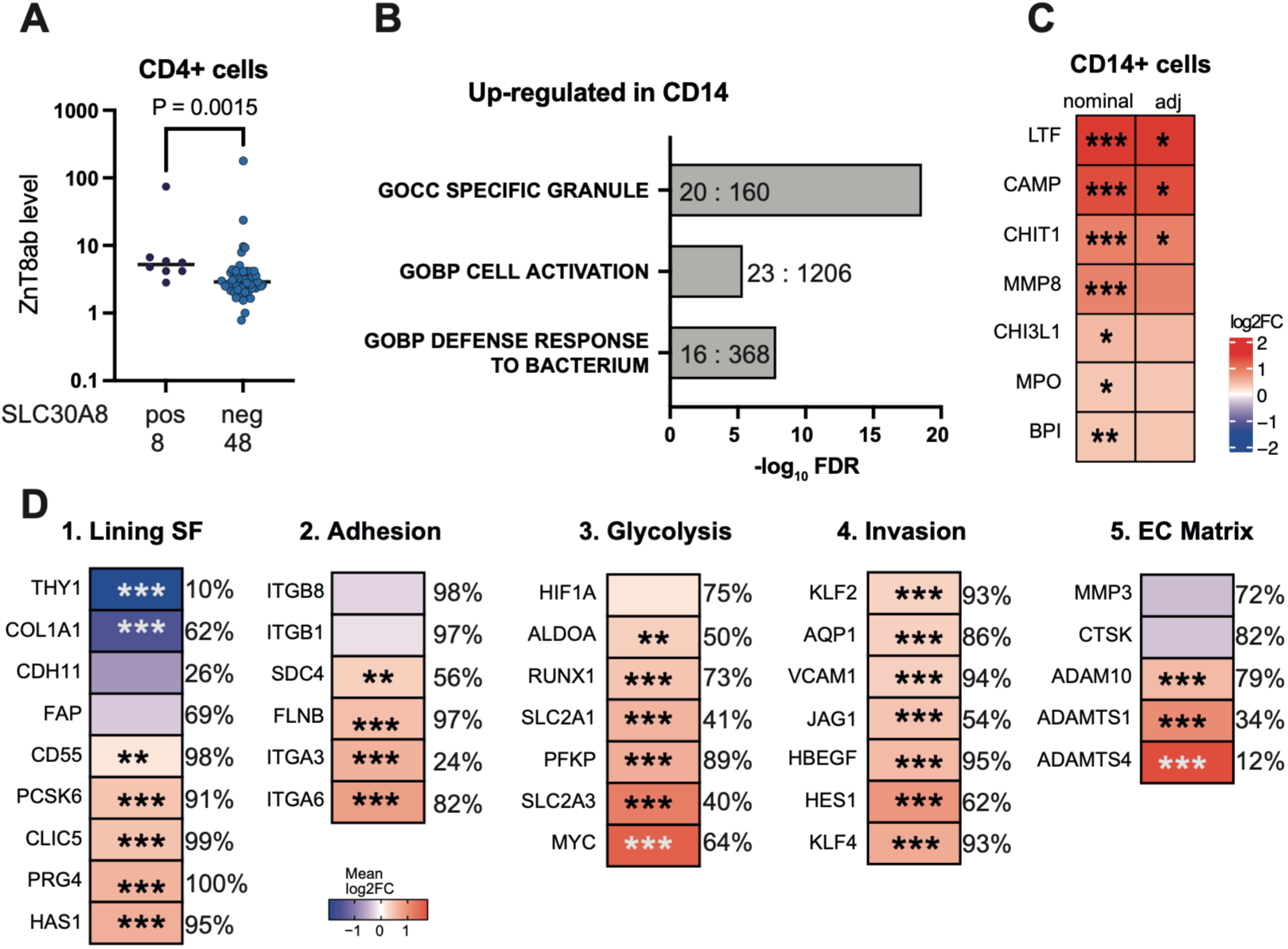
Expression of ZnT8 gene SLC30A8 in CD4+ T cells and synovial tissue of patients with rheumatoid arthritis. A. Dot plot of ZnT8ab levels in serum of 56 female RA patients grouped by expression of SLC30A8 gene in CD4+ cells. Expression was measured by RNA sequencing. Positive group had the normalized expression above 1. B. Bar plot of the top biological pathways enriched by expression of genes upregulated in CD14+ cells of 56 female RA patients in regression to increasing serum levels of ZnT8Aab. 194 upregulated protein coding genes with normalized expression >10 and nominal p-values <0.05 identified by DEseq2, were included in the enrichment analysis. Number of upregulated to the total gene number within each pathway is showed inside bars. C. Heatmap of relative expression difference (by log2FC) for the top genes upregulated in CD14+ cells in regression to serum levels of ZnT8ab. P-values <0.05 (*), <0.001 (**), <0.0001 (***). D. Heatmap of relative expression difference (by log2FC) of selected genes in SLC30A8+ cells in comparison to the remaining cells within the lining synovial fibroblast cluster grouped by function. The normalized gene expression is calculated in single-cell transcriptome of RA synovial tissue (Synapse, syn52297840). The expressing cell frequency among the synovial stromal cells is shown as %.

Analysing transcriptome of blood CD14^+^ cells in regression to serum Znt8ab levels, we found that increasing levels of ZnT8ab were associated with marked changes in the transcriptome of CD14^+^ cells. These changes were reflected by a) enrichment for secretory vesicles (GO:0042581, 20/159, FDR 4.47e-12) through a significant upregulation of *LTF/*lactoferrin, *CAMP*/LL37, *CHI3L1*/YKL40, *BPI*, *MPO* genes; b) upregulation of matrix remodelling program genes *MMP8, ECM1, ANXA3, CLU, CPAMD8* (**Figure 5B, 5C**). In contrast to CD4^+^ cell, none of CD14^+^ cell subset had *SLC30A8* expression levels above 1. These bioinformatic findings implied enrichment for cells loaded with enzymes, oxidant-generating machinery, and granule proteins capable of damaging the extracellular matrix and surrounding tissue. Together, this embodies a particular phenotype of CD14^+^ cells activated by danger signals and ready for defence against endogenous or external insults. This CD14^+^ cell phenotype was enriched in direct proportion to ZnT8ab production in non-diabetic RA patients.

Collectively, these findings provide evidence that ZnT8ab levels were associated with an activated secretory type of CD14^+^ cells and suggest that ZnT8 antigen exposure outside the pancreas could trigger ZnT8ab production in RA. To investigate this possibility, we next analysed the expression of ZnT8 in synovial tissue of RA patients.

### 5. Expression of ZnT8 is annotated to the lining layer of synovial fibroblasts

Single cell transcriptomic data from synovial tissue (ST) biopsies of 82 RA patients ^34^ were retrieved and subclusters of T cells (24 clusters), myeloid cells (15 clusters), stromal cells (11 clusters) were investigated for *SLC30A8* expression coding for ZnT8A. This analysis revealed that stromal cells contained the largest subset of *SLC30A8* gene expressing cells coding for ZnT8, followed by T cells, although the overall numbers of *SLC30A8*^+^ cells were low (**Table 3**). Focusing on the ST stromal cells, we found *SLC30A8*^+^ cells to be located exclusively among the lubricin-producing *PRG4*^+^ synovial fibroblasts (SF) of the lining layer, with a normalized expression level of 5.6 nCPM in the *PRG4*^+^ cell cluster. The abundance analysis of the *SLC30A8*^+^ cells in comparison to the remaining synovial lining cells revealed expression of the key gene characteristic for *PRG4*^+^*CLIC5*^+^ lining synovial fibroblasts in >92% of cells (**Figure 5D1**). Consistent with the lining fibroblast phenotype, *SLC30A8*^+^ SF expressed low levels of *THY1*/CD90, *PDPN*, and *CDH11*. Therefore, the lining *SLC30A8*^+^ SF were adhesion-competent cells programmed for matrix interaction and expressing *ITGA6* and *ITGA3* combined with *ITGB1* form the laminin-oriented adhesion integrin pairs α3β1 and α6β1, that navigate and anchor the leading edge of ST and pannus growth (**Figure 5D2**).

**Table 3.** Expression of the *SLC30A8* gene coding for ZnT8 in RA synovial tissue. *Ad hoc* analysis of public single-cell RNAseq, Zhang Nature 2023.

| Cell type | Total cells | Expressing SCL30A8, n (%) | Mean expression |
| --- | --- | --- | --- |
| Stromal cell | 79555 | 454 (0.57) | 0.58 |
| T cell | 94048 | 20 (0.02) | 1.04 |
| Myeloid cell | 76171 | 8 (0.01) | 0.53 |
| B/plasma cell | 30697 | 4 (0.01) | 0.38 |
| NK | 8497 | 4 (0.05) | 0.91 |
| Endothelial cell | 25043 | 1 (0) | 0.71 |

The *SLC30A8*^+^ SF displayed a glycolytic shift of metabolism expressing high levels of glucose transporters GLUT1/*SLC2A1* and GLUT3/*SLC2A3*, and the genes of *MYC*, *PFKP* and *ALDOA* which maintained the glycolysis commitment of these cells (**Figure 5D3)**. This glycolytic shift fuelled cell migration and invasive growth executed by the *HBEGF, EREG, AQP1*, *ANXA3, ADAM10, SDC4*, and *FLNB* genes dependent on the Wnt, NOTCH, and AP1 signalling pathways, all of which were over-represented in *SLC30A8*^+^ SF (**Figure 5D5)**. Despite activation of the Notch and AP1 signal, *SLC30A8*^+^ cells were not fully mature pannus cells since the expression of *CDH11*, *MMP3* and *OPG/TNFRSF11B* essential for cartilage and bone invasion, remained low.

Taken together, the single-cell transcriptome of the lining *SLC30A8*^+^ SF revealed adhesion-competent cells programmed for matrix interaction and invasive behaviour. These cells may therefore present a natural elimination target in RA, potentially justifying ZnT8ab production.

### 6. Expression of the *SLC30A8* gene in RA synovial fibroblast cultures

To investigate whether inflammation or insulin acts as a positive contributor to ZnT8 expression in human synovial fibroblasts, we used 16 independent SF cultures across three independent experiments. RASF cultures were stimulated with TNF or IL1β to induce inflammation, which was confirmed by the increased IL-6 cytokine levels in the culture supernatants (**Figure 6A**). RASF activation during 24h by neither TNF nor IL1β was associated with an increase in *SLC30A8* expression as measured using conventional RT-PCR, although the mRNA levels were overall low (**Figure 6A**). Next, RASF cultures (n=8) were treated with a combination of TNF and insulin for 24h and 96h. Response to insulin was clearly measurable in RASF by a suppression of the intracellular insulin mediator *IRS2* and by downregulation of IL-6 production (**Figure 6A and 6B**). SF treated with insulin for 96h upregulated significantly *SLC30A8* expression (p=0.026) (**Figure 6C**). Taken together, these results showed that RASF express low but measurable levels of *SLC30A8* that were also responsive to insulin stimulation.

**Figure 6.**
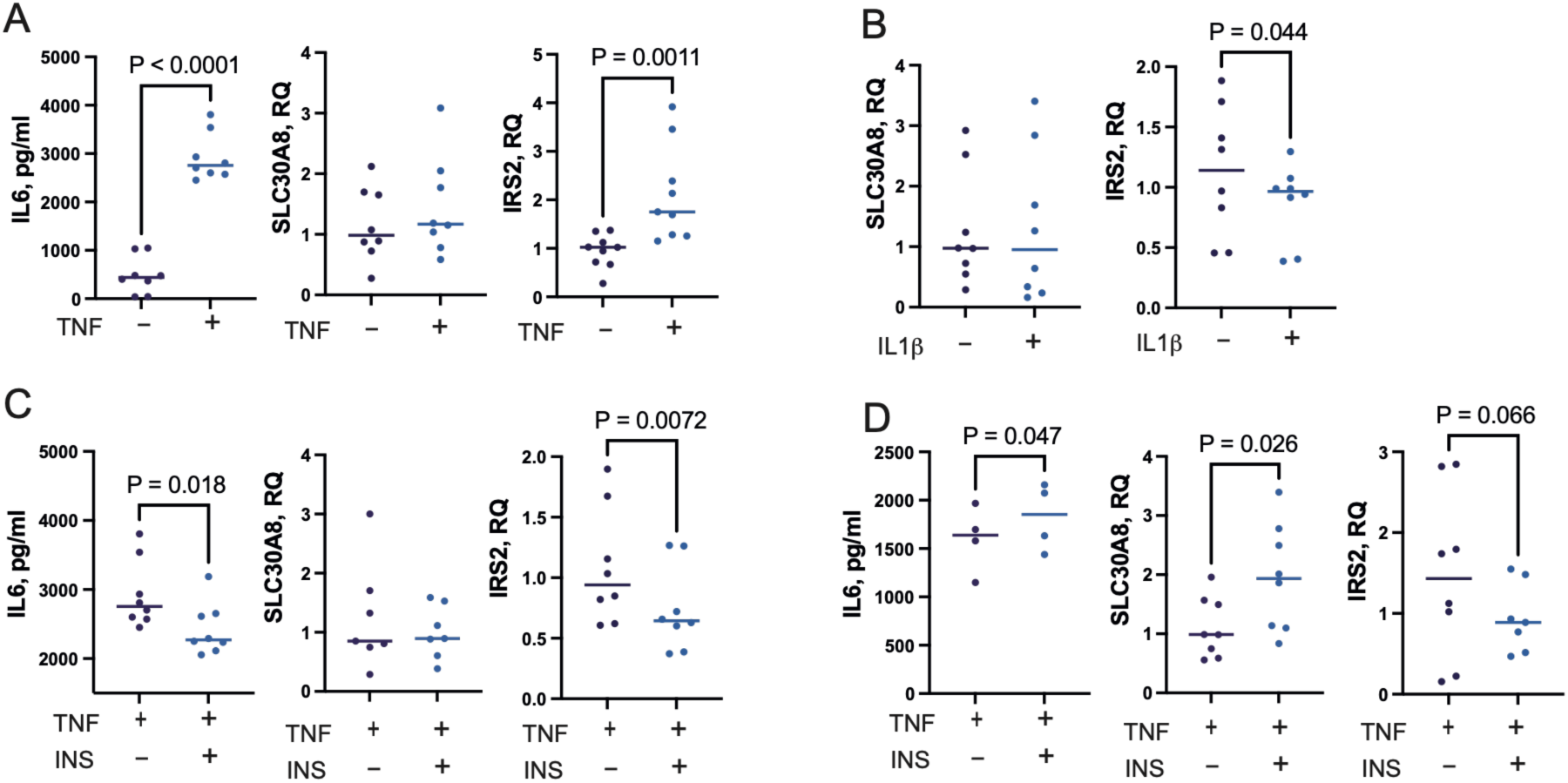
Expression of SCL30A8 coding for ZnT8 in synovial fibroblast cultures of patients with rheumatoid arthritis. A, B. Eight independent synovial fibroblast cultures were prepared from knee synovial tissue and stimulated with (A) TNF (10ng/ml for 24h) or (B) IL1b (10ng/ml for 16h). Gene expression was measured by RT-PCR, IL6 levels were measured in supernatants by an ELISA. C, D. Eight independent fibroblast cultures stimulated with TNF was also treated with a combination of TNF and insulin (both, 25 nM) for 24h and 96h. Insulin was added daily. Gene expression was measured by RT-PCR, IL6 levels were measured by an ELISA. P-values are calculated by Student’s paired t-test.

## Discussion

This study revealed a frequent production of DMab in non-diabetic RA patients, detected in 54% of RA patients, and predominantly in women. The DMab profile in RA patients was dominated by the reactivity against ZnT8 which differed from T1D, LADA, and other autoimmune conditions, where GAD65ab were most detected ^1, 2, 3^). Our recent 10-year follow-up of RA patients demonstrated that DMab were not accompanied by the development of DM ^9^. This prompted us to investigate whether DMab, as part of autoimmune continuum, may reflect a broader autoimmune activation with biological relevance for RA and systemic immune dysregulation.

Guided by the intention to increase sensitivity for early detectable autoimmune responses, we applied a low positive cut-off for DMab measurements and provided here an evidence of their biological relevance in RA. In absence of clinically obvious DM, ZnT8ab production was still connected to an impaired glucose metabolism as reflected by higher insulin levels, higher levels of glycated haemoglobin (HbA1c), and a higher HOMA-IR index. Additionally, anabolic factors including insulin and BDNF were identified as potential triggers of Znt8ab production providing a mechanistic link to the glycolytic shift observed in ZnT8/*SLC30A8*^+^ fibroblasts in RA synovial tissue. Both, insulin and BDNF, are known to exert context dependent anti-inflammatory effects, including inhibition of cytokine production and anti-inflammatory polarization of T cells and macrophages ^10, 11, 35, 36, 37^. Accordingly, we found a tight association between ZnT8ab and a distinct serological profile characterized by low serum levels of pro-inflammatory cytokines IL-6, IFNg and IL-9 in non-diabetic RA patients. This differs from patients with T2D and obesity, in whom hyperinsulinemia is generally associated with a pro-inflammatory state and may be treated by anti-cytokine approaches ^38, 39, 40, 41^. In addition to this anti-inflammatory environment, the increasing ZnT8ab levels were produced in direct proportion to the ZnT8 gene expression in CD4^+^ cells. Although the biological significance of this finding is uncertain because of barely detectable amounts of *SLC30A8* transcripts, the association denotes that antigen exposure outside the pancreas could contribute to ZnT8ab production in RA patients. This finds support in ZnT8 expression within lubricin-producing cells of the lining SF layer of RA joint tissues. Increasing ZnT8ab levels were also associated with enrichment for activated CD14^+^ macrophages loaded with secretory lectins and granule-associated mediators including lactoferrin, LL37, YKL40, and MPO. These factors trigger immediate extracellular neutralisation of target or cover the macrophage extracellular traps thereby enhancing their efficacy ^42^. Together, these evidenced for the functional relevance of ZnT8ab and imply that Znt8ab production in RA was associated with low inflammatory state supporting resolution of inflammation. We further found that the increasing levels of ZnT8ab significantly reduced the probability of RAab-production in the studied patients. This points at independent triggering mechanisms behind production of DMab and RAab in RA patients advocating for their different biological roles in RA pathology. The observation that non-overlapping variables were associated with production of DMab and RAab in regression models further strengthened this notion.

We established that in RA synovial tissue, expression of *SLC30A8* was almost solely attributed to the *PRG4*^+^ lubricin-producing SF, which formed a thin lining layer barrier between the joint cavity and the underlying tissue ^43, 44, 45^. In RA, the lining SF undergo a tumour-like transformation, becoming aggressive, proliferating, and pannus-forming tissue above the cartilage causing its damage ^46, 47^. All these features were prominent in the *SLC30A8*^+^ SF cluster in the analysed RA tissue, which transcriptome was shaped by hypoxia and shear stress and epigenetic reprogramming through the glycolytic switch. Nevertheless, the lining SF lack a basement membrane which makes them vulnerable for autoreactive cells and autoantibodies enriched in the joint compartment. Thus, our findings support a scenario where *SLC30A8*^+^ SF of the lining layer are targeted by the damage-primed CD14^+^ cells and ZnT8ab.

According to the single-cell summary presented by the Human Protein Atlas, ZnT8 gene SLC30A8 is highly expressed in pancreatic islet cells (665 nCPM) and in renal nephron epithelial cells (132 nCPM). In contrast, production of ZnT8 in acinar cells of eye (13.6 nCPM), salivary gland (3.3 nCPM) and pancreas (4.1 nCPM), which share exocrine functions, was well comparable to the one found in the lining SF (5.1 nCPM) and was substantially higher compared to blood T cells. The importance of zinc transferring proteins for joint homeostasis has been studied in experimental mouse models ^48^. Zinc transporters have been shown vital for lubricin maturation and storage in exosome vesicles ^45, 49^. Deficiency in zinc importer SLC39A8 is implicated in cartilage degeneration ^50^. Lubricin-producing cells in the joints, eyes, and the lungs form protecting anti-adhesive, and anti-inflammatory surfaces, where lubricin may functionally cooperate with ZnT8. In this context, the reported high frequency of ZnT8ab in patients with autoimmune uveitis ^8^ advocates for a zinc transport dependent bridge between joint and ocular autoimmune pathology.

Taken together, this study demonstrates that zinc transporter ZnT8 is a novel antigenic target in RA. Expression of ZnT8 in the lining SF of RA synovial tissue encourage the biological importance of ZnT8ab for arthritis pathology. Our study suggests that ZnT8ab in RA should not be interpreted solely as preclinical marker of autoimmune diabetes. Instead, they may represent a non-canonical, tissue-context-dependent autoantibody response, in which ZnT8 antigen exposure occurs outside the pancreatic islet and is linked to synovial tissue remodelling rather than progression to diabetes. These findings shed new light on understanding of RA heterogeneity where DMab and RAab identify biologically distinct autoimmune trajectories within RA rather than representing overlapping manifestations of generalized autoimmunity.

Some limitations of this study should be acknowledged. First, *SLC30A8* expression outside pancreatic islets was low at the transcript level, and its biological interpretation requires caution. Second, localization of *SLC30A8* expression in RA synovial tissue was based on single-cell transcriptomic data and should be validated at the protein level in independent RA tissue cohorts. Third, although the association between ZnT8ab, *SLC30A8*^+^ lining synovial fibroblasts, and activated CD14^+^ cells supports a biologically plausible model, the present study does not directly demonstrate binding of ZnT8ab to synovial fibroblasts or antibody-mediated elimination of these cells. Future studies should address whether ZnT8ab recognize ZnT8 protein in RA synovium and whether this response modifies fibroblast survival, macrophage activation, or pannus progression.

## Acknowledgements

We would like to thank the research nurses Anneli Lund and Marie-Louise Andersson at the Rheumatology Clinic, Sahlgrenska University Hospital, Gothenburg, for their help with blood sampling. We are thankful to Dr. Anna-Karin Huldgård Ekwall, Rheumatology Clinic, Sahlgrenska University Hospital, for sharing synovial fibroblast cultures. We also thank all patients with RA who participated in this study.

## Author contribution

Patient material, MIB, RP, CM, EN, UML; Data acquisition and analysis, MCE, VC, EN, MYHA, YZ, STS, RP, MIB; Visualization, MCE, VC; Writing original draft, MIB, RP; Review & editing, MCE, VC, EN, STS, CM, UML, RP, MIB; Project administration, STS; Funding acquisition, RP, MIB; Conceptualization and supervision, MIB.

## Data Availability

Transcriptional data are available as Fastq-files and processed reads, deposited in Gene Expression Omnibus (GEO) at the National Centre for Biotechnology Information with the accession codes GSE201669 and GSE201670

## Supplementary material

**Supplementary Figure S1.**
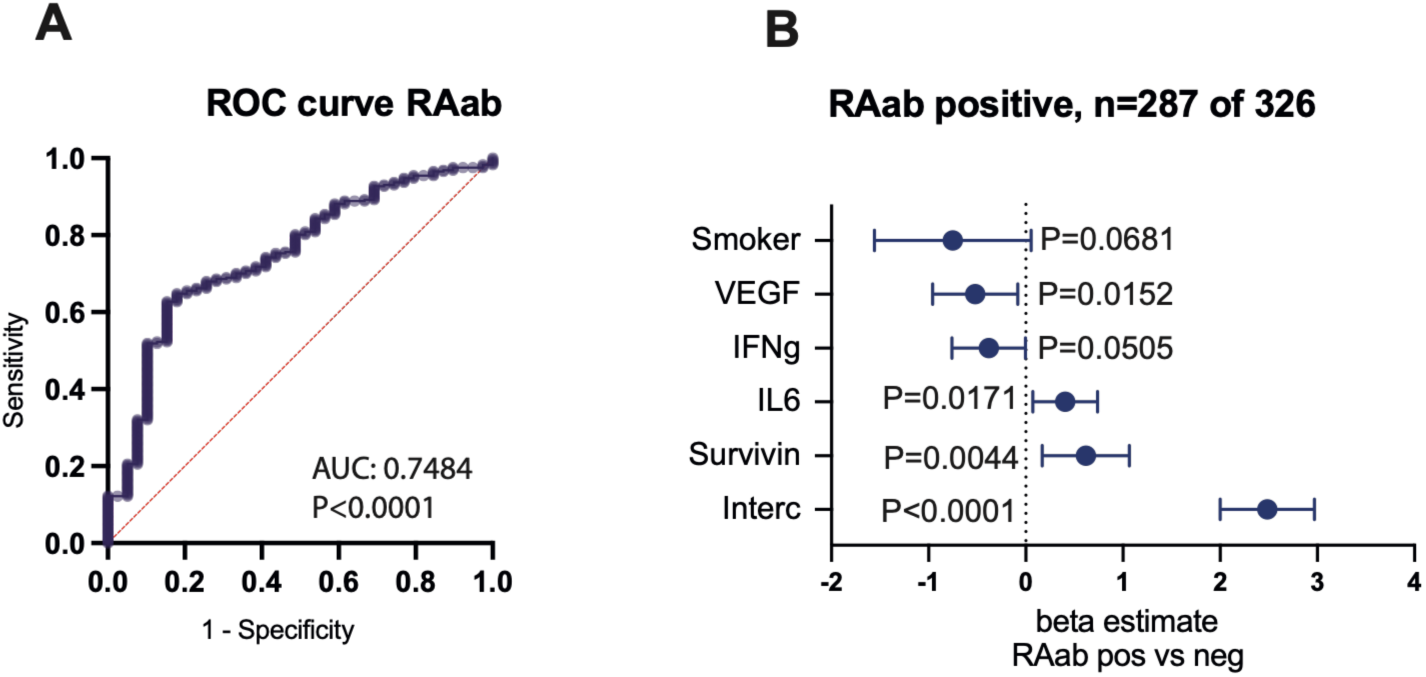
Serological associations of rheumatoid arthritis-related antibodies (RAab) in patients with rheumatoid arthritis. RAab to cyclic citrullinated peptides and rheumatoid factor were measured in 326 non-diabetic RA patients. In total, 287 patients produced any or both of RAab. A. Receiver Operating Characteristic (ROC) curve of prediction for RAab positivity was calculated in the binary multivariate logistic regression analysis using presence of RAab as dependent to 30 serological and clinical variables. Area under the ROC curve (AUC) was calculated to estimate the accuracy of the model. B. Forest plot of beta estimates for the variables included in the RAab prediction model. Dots indicate the beta estimate values, whiskers indicate 95% confidential interval. P-values are calculated by Wald statistics.

**Supplementary figure S2.**
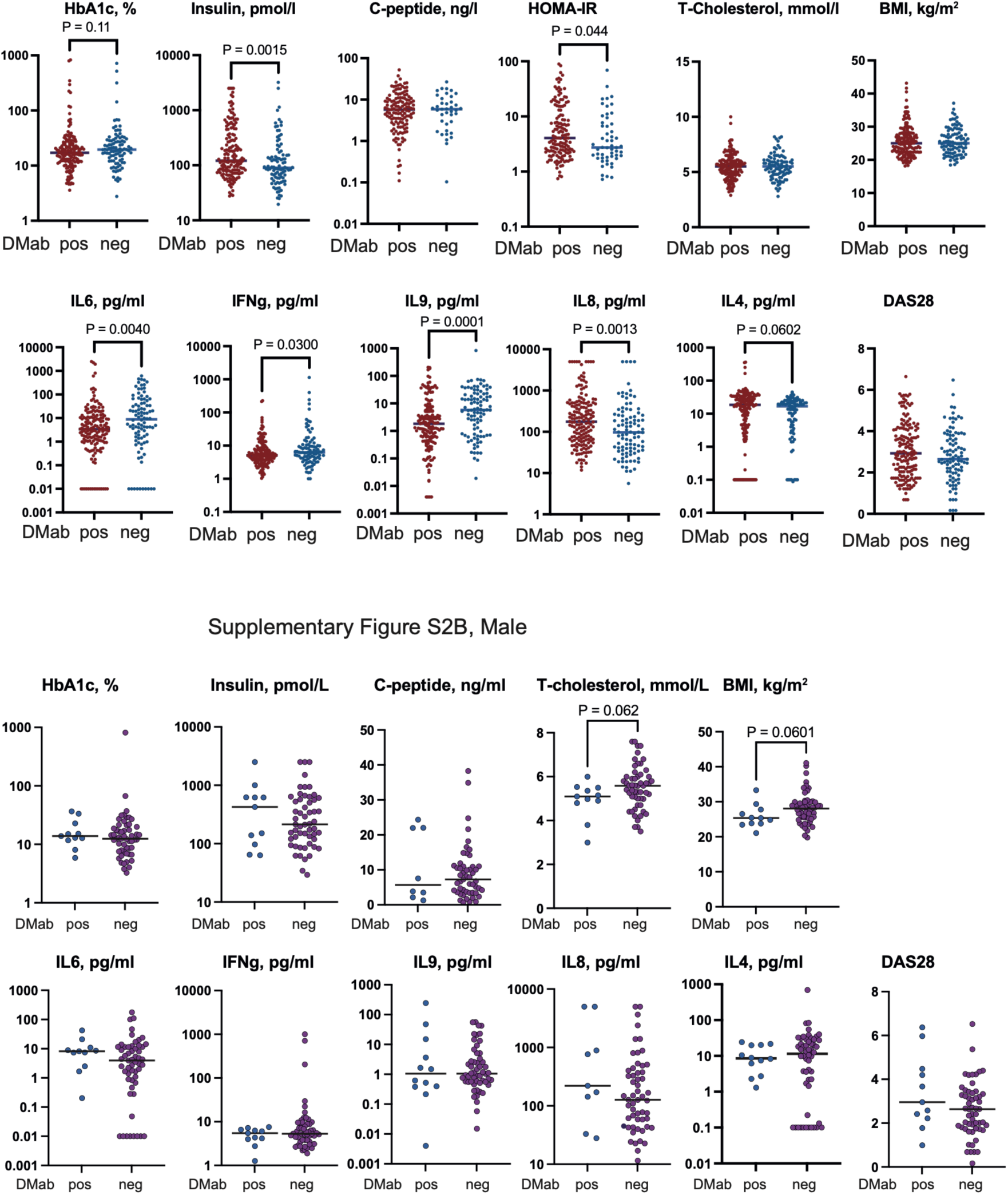
Serological and clinical characteristics of non-diabetic female (A) and male (B) patients with rheumatoid arthritis grouped as DMab positive vs negative. Horizontal line indicates median for group. Group comparisons were done pairwise using non-parametric Mann-Whitney U test.

**Supplementary Figure S3.**
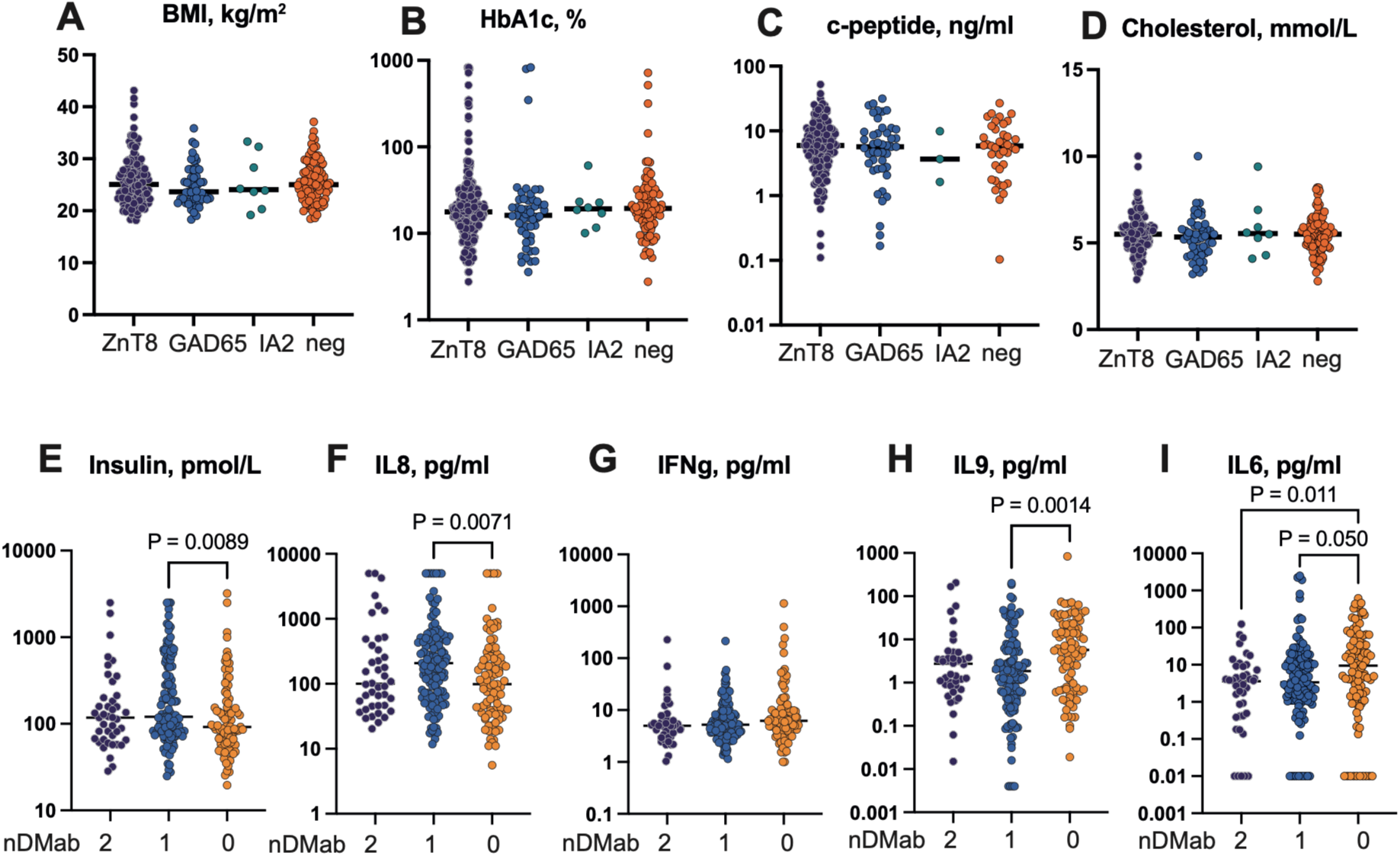
Serological and clinical characteristic of non-diabetic female patients with rheumatoid arthritis grouped by the presence of different type (A, B, C, D) and the number (E, F, G, H, I) of diabetes mellitus-related antibody (DMab). DMab including ZnT8ab, GADab, IA2ab were measured in serum of 260 female RA patients. Patients positive for several antibodies are shown within each group. Individual variables are presented by dot plots where horizontal line indicates median for group. Group comparisons were done pairwise using non-parametric Mann-Whitney U test.

**Supplementary figure S4.**
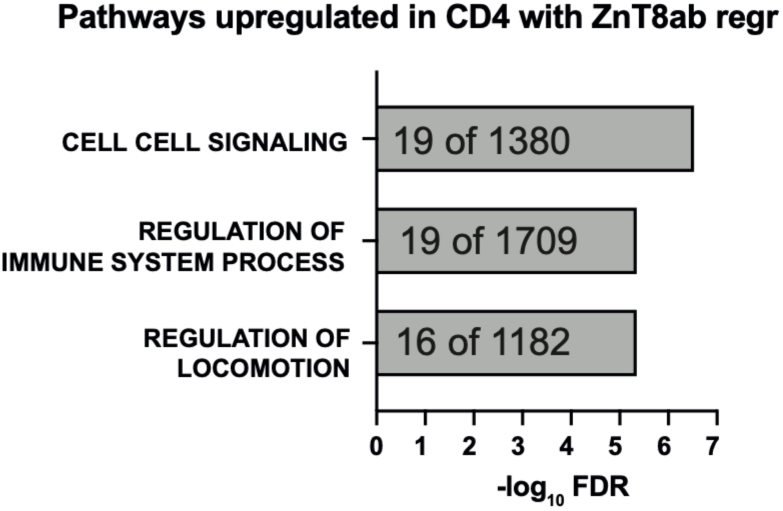
Bar plot of the top biological pathways enriched by expression of genes upregulated in CD14+ cells of 56 female RA patients in regression to increasing serum levels of ZnT8Aab. 194 upregulated protein coding genes with normalized expression >10 and nominal p-values <0.05 identified by DEseq2, were included in the enrichment analysis. Number of upregulated to the total gene number within each pathway is showed inside bars.

**Supplementary table S1.**
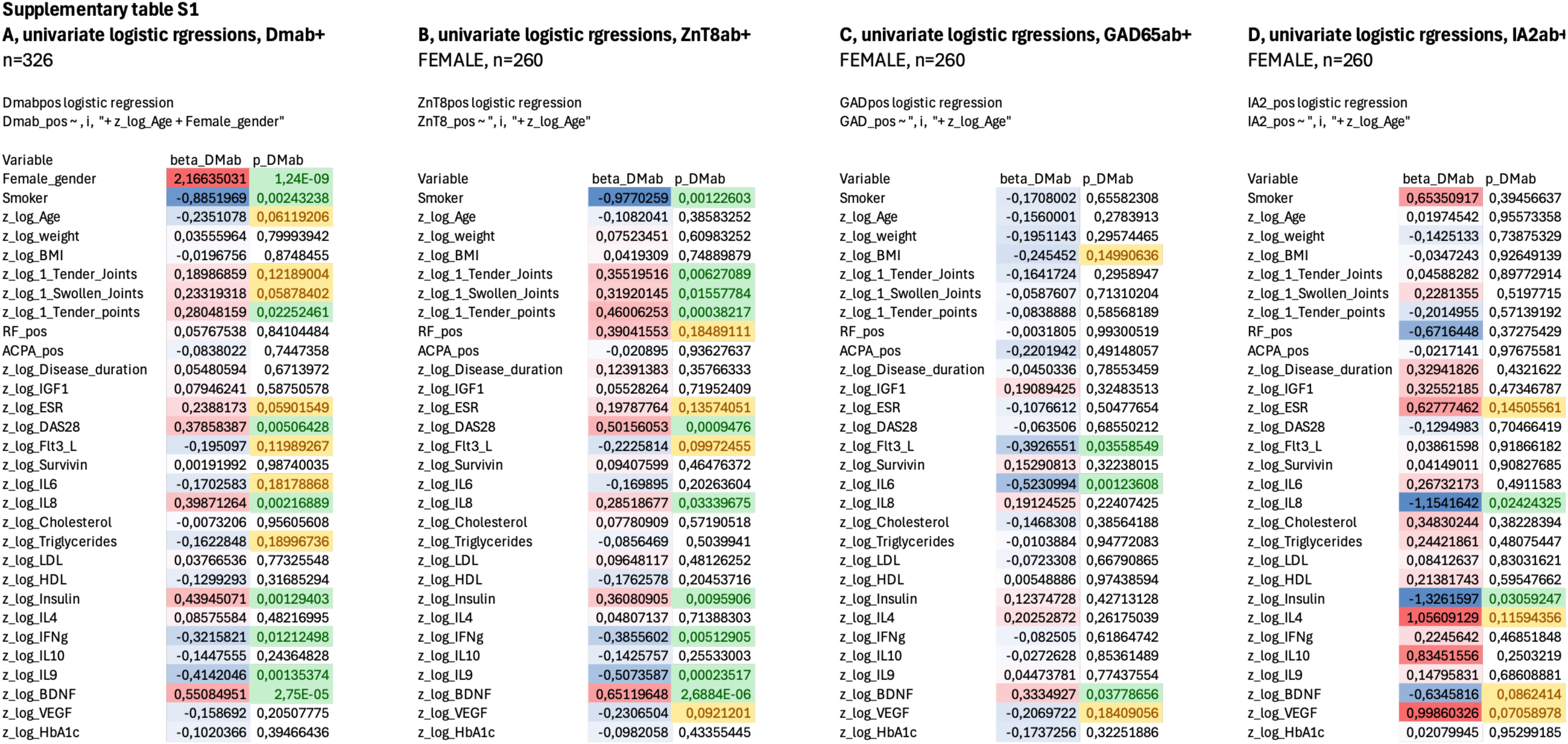

